# The prevalence of common and stress-related mental health disorders in healthcare workers based in pandemic-affected hospitals: a rapid systematic review and meta-analysis

**DOI:** 10.1101/2020.05.04.20089862

**Authors:** Sophie M. Allan, Rebecca Bealey, Jennifer Birch, Toby Cushing, Sheryl Parke, Georgina Sergi, Michael Bloomfield, Richard Meiser-Stedman

**Author notes:** Corresponding author: Richard Meiser-Stedman, Department of Clinical Psychology and Psychological Therapies, Norwich Medical School, University of East Anglia, Norwich Research Park, Norwich, NR4 7TJ, UK.

## Abstract

**Background:** Healthcare workers (HCWs) are considered at elevated risk of experiencing mental health disorders in working with patients with COVID-19.

**Aims:** To estimate the prevalence of common mental health disorders in HCWs based in hospitals where pandemic-affected patients were treated.

**Method:** Databases were searched for studies published before 30th March 2020. Quantitative synthesis was used to obtain estimates of the prevalence of mental health disorders in four time windows, determined a priori (the acute phase, i.e. during and up to 1.5 months post-pandemic; 1.5-5.9 months; 6-11.9 months; 12 months and later).

**Results:** Nineteen studies met the review criteria. They predominantly addressed the acute phase of the SARS outbreak in Asia. The most studied outcomes were clinically-significant post-traumatic stress symptoms (PTSS) and general psychiatric caseness. For clinically significant PTSS in the acute phase, the prevalence estimate was 23.4% (95% CI 16.3, 31.2; N=4147; I2=96.2%); in the 12 months plus window, the estimate was 11.9% (8.4, 15.8; N=1136; I2=74.3%). For general psychiatric caseness, prevalence estimates were: acute phase, 34.1% (18.7, 51.4; N=3971; I2=99.1%); 6-12 months, 17.9% (13.1, 23.2; N=223; I2=0.0%); 12 months plus, 29.3% (6.0, 61.0; N=710; I2=97.8%).

**Conclusions:** Mental health disorders are particularly common in HCWs working with pandemic-afflicted patients immediately following a pandemic, but the course of disorders following this period is poorly understood. PTSS remained elevated compared to the general population at 12 months, despite there being some evidence for natural recovery. There was considerable heterogeneity, likely linked to methodological differences. More extended follow up of HCWs is needed.

## Introduction

Healthcare workers (HCWs) including nurses, doctors, allied health professionals and all support staff based in hospitals where patients with COVID-19 are treated face considerable challenges and stress. In addition to the clinical challenges associated with treating a large volume of severely unwell patients, HCWs working with this group of infectious patients face threats to their own physical health^1^, with a number of highly publicised HCW deaths already reported due to COVID-19. There is increasing recognition of the significant psychological impact of caring for those with COVID-19 given the immense pressure facing HCWs. For example, HCWs may face situations where they are at risk of sustaining moral injury^2^, while there are also difficulties in obtaining sufficient personal protective equipment.^3^ Health systems are subsequently implementing mental health provision systems and additional psychological support^4^. In order to better plan and develop these support systems, and to assist with education around reactions to working with COVID-19 patients, a rapid systematic review was undertaken to determine the prevalence of mental health disorders in HCWs working with patients infected through a pandemic. We broadened our search to include studies relevant to the current COVID-19 crisis, e.g. pertaining to other coronavirus outbreaks (SARS, MERS) and other epidemics that represent significant risks to HCWs (e.g. Ebola). In particular we sought to establish the prevalence of different conditions at different phases, i.e. during and immediately after a pandemic, then over the following months.

## Method

### Protocol and registration

The present review was not registered given the perceived need to disseminate a rapid review pertaining to the mental health consequences for HCWs given the exponential rise in hospital admissions and deaths for COVID-19. The review was produced in accordance with Preferred Reporting Items for Systematic Reviews and Meta-analysis (PRISMA) recommendations^5^.

### Eligibility criteria

Studies were included in the present review if they measured the prevalence of mental health disorders (in particular clinically significant post-traumatic stress, depression, anxiety or general psychopathology) in healthcare workers that worked in a hospital where care was provided to patients who had acquired an infection because of a pandemic, e.g. SARS, MERS, Ebola, COVID-19. No restrictions were placed on healthcare worker type (e.g. medical and non-medical staff were included) or department worked in.

Studies, or partial study data, were excluded if they; i) focused exclusively on healthcare workers who had also developed the index illness at the centre of the pandemic; ii) the sample included in-patients with the index infection; iii) the study was not published in English; iv) the study reported on stress or occupational wellbeing measures, such as burnout, rather than a diagnosable or clinical significant mental health disorder; v) participants included staff at other non-affected hospitals; vi) only comprised qualitative data; or vii) addressed work with the HIV pandemic.

### Information source

Databases searched included Medline, PsycINFO, CINAHL, PubMed, OVID and ScienceDirect. Manual searches of relevant review papers and empirical articles were also carried out to identify any studies that had not yet been included in the literature databases.

### Search

Search terms were 1) terms related to identified pandemics (including SARS, MERS, Coronavirus, Ebola and ‘pandemic’) AND 2) ‘acute hospital (including all search engine variants) AND 3) ‘mental health’ (including post traumatic stress, depression, anxiety and low mood variants) AND 4) ‘health^*^ professional’ (including variants such as doctor and nurse). Searches were conducted on 30th March 2020. See Supplementary Material 1 for full search terms.

### Data collection process

Duplicates were removed from search results. Titles and abstracts were screened for eligibility. The full texts of eligible studies were then accessed and checked against the inclusion and exclusion criteria. Six researchers were split into pairs (SA and RB, JB and SP, and TC and GS), both of whom independently completed initial screening and data extraction. Disagreements were resolved by discussion with the wider team and a decision reached by consensus.

### Data items

Descriptive data was extracted pertaining to key study characteristics (country and year of publication, pandemic, sex and role of participants, method of data collection). Data from comparison control groups (e.g. HCWs at another hospital that did not work with pandemic patients) were not extracted. Prevalence data were extracted for post-traumatic stress symptoms (PTSS), depression, anxiety and general psychiatric screening using the number of participants who scored above a defined cut-off on the given outcome measure or met threshold for a diagnosis based on a structured interview. Data were categorised according to four time periods, which were defined *a priori:* during the pandemic up to 1.49 months later (termed the ‘acute phase’); 1.5-5.9 months; 6-11.9 months; and 12 months or later.

### Risk of bias

The quality of the included papers was assessed using an adapted version of the National Heart, Lung and Blood Institute Quality Assessment Tool for Observational Cohort and Cross-Sectional Studies (https://www.nhlbi.nih.gov/health-topics/study-quality-assessment-tools; see Supplementary Material 2 for full rating scheme). The assessment criteria included: study population defined; participation rate of over 50%; if follow-up, was the attrition rate described; and validity and reliability of measures for post traumatic stress, anxiety, depression and general psychiatric screen. Individual studies were scored for quality (0= not present/poor description, 1= some description but some missing information, 2= all desired information included) for each area of quality. Studies were given a percentage according to the degree of criteria being met. The percentage was used to indicate the study quality (>70% high, 50-69% medium, <50% low).

### Quality assessment and extraction were double rated; there were no disagreements on quality rating

Given how few studies were included in each meta-analysis (less than the 10 suggested for funnel plot asymmetry)^6^, formal tests of publication bias are not reported.

### Summary measures

The summary measure of interest was prevalence of a mental health difficulty, based on number scoring above cut-off on a self-report questionnaire measure (i.e. clinically significant levels of symptoms) or the number meeting diagnostic threshold based on a structured interview.

### Synthesis of results

Prevalence outcomes were synthesised using a random-effects meta-analysis. Arcsine transformations was used to account for issues with study weightings when estimating prevalence^7^, with back-transformed values presented in all results. The metafor package^8^ in R 3.4.2^9^ was used to conduct the meta-analysis.

## Results

### Study selection

The numbers of studies screened, assessed for eligibility, and included in the review, with reasons for exclusions at each stage, are presented in a PRISMA flowchart (see Figure 1). Nineteen studies provided usable data. Two articles related to the same cohort but at different time intervals^10 11^; these are therefore reported as a single study.

**Figure 1.**
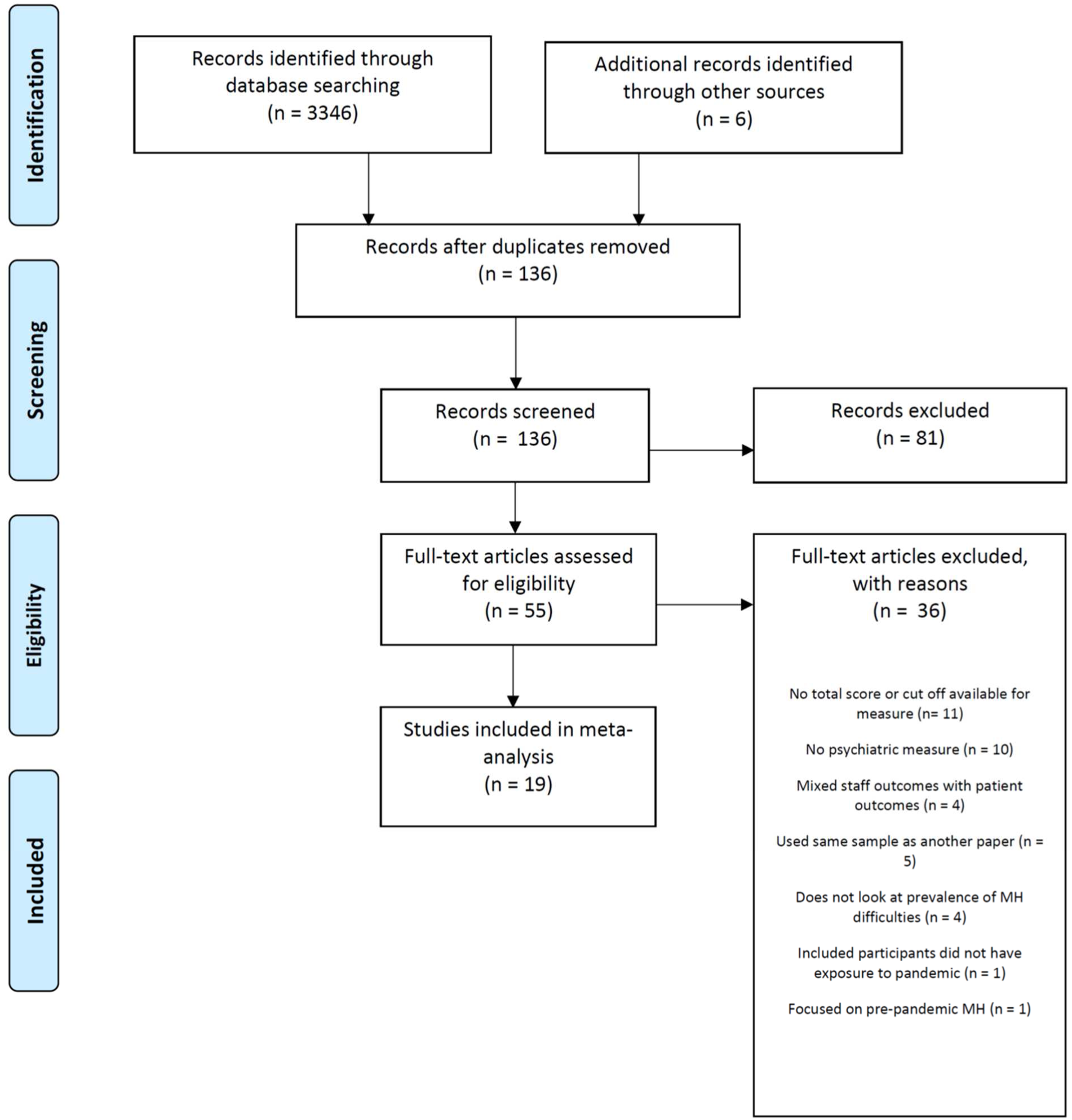
PRISMA flowchart.

One group reported two articles on the same hospital but the sampling frame for the follow up study was so different to the first that these articles are reported separately^12 13^. The same Canadian study group also reported follow up data on a smaller sub-set of participants but using structured interviews rather than self-report questionnaire screening^14^; a separate note on this additional study is provided below. References for all articles included in the review are provided in Supplementary Material 3.

### Study characteristics

Study characteristics are reported Table 1. The majority of included studies related to the SARS pandemic in Asian hospitals. One study reported findings from the COVID-19 pandemic. One set of studies pertained to the SARS outbreak in Canada, and a further study pertained to the H1N1 pandemic in Greece. All but two studies involved mixed healthcare worker samples; the remaining two focused exclusively on nurses. Reported outcomes were classified as being related to PTSS, depression, anxiety or a general psychiatric screen. Three studies reported that mental health interventions were offered to HCWs in response to the pandemic.

**Table 1.**
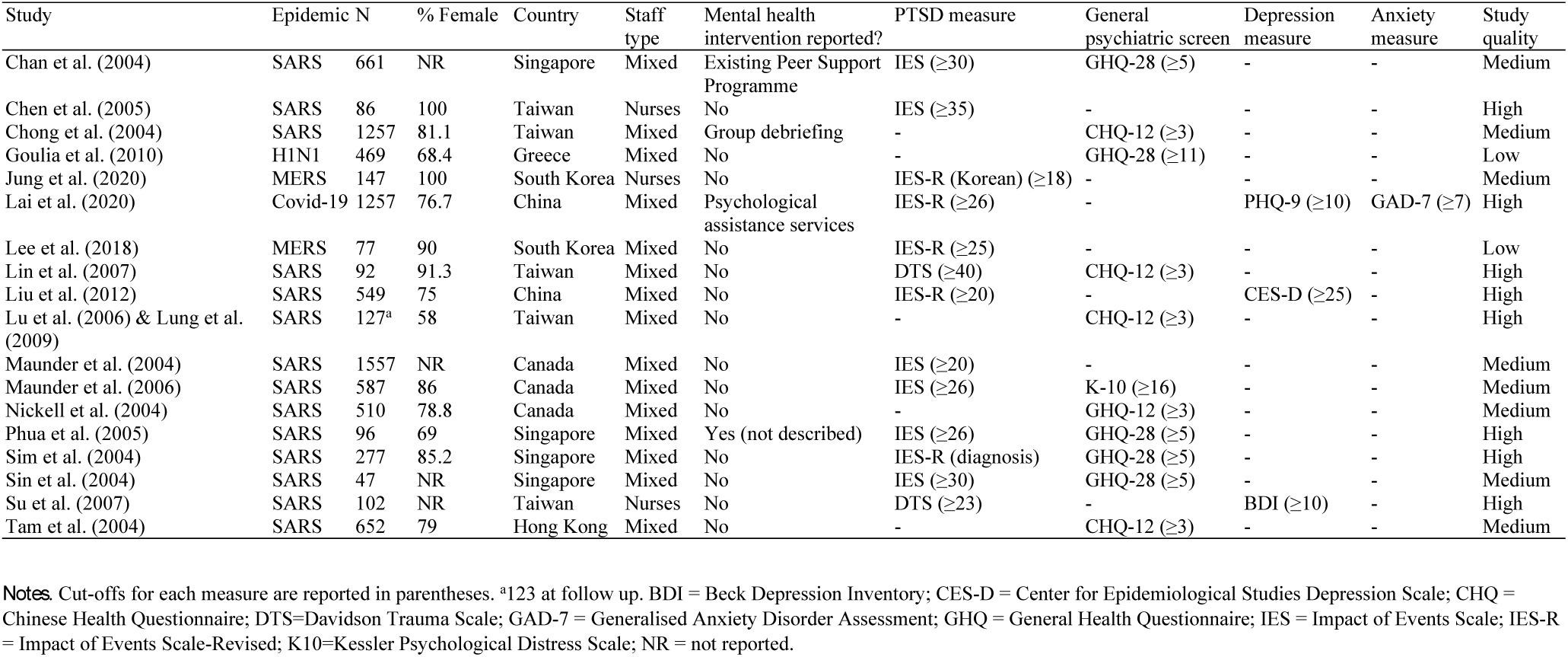
Study characteristics.

### Risk of bias within studies

Overall, eight studies were rated as high quality, eight as medium quality and two as low quality (Table 1; see Supplementary Material 4 for full quality ratings). The majority of studies (15 of 18) were judged to have clearly specified their study population. Ten of the studies had a participation rate greater than 50%. All assessment tools were self-report questionnaire measures; one study used a structured interview with a sub-set of HCWs, which will be reported separately. In only two studies was it explicitly stated that a PTSS measure was completed in relation to the relevant pandemic.

### Prevalence of psychiatric disorders

**Prevalence of clinically significant post-traumatic stress symptoms (PTSS)**. Forest plots for each time window are displayed in Figure 2. The pooled estimate for clinically significant PTSS in the acute phase (i.e. during the pandemic itself and up to 1.5 months after the end of the pandemic) was 23.4% (95% CI, 16.3, 31.2; k=9; N=4147). The pooled studies had a large degree of heterogeneity (Q[8] = 190.00, p < .0001; I^2^=96.2%). A single study addressed the 1.5-5.9m window (57.1%; 95% CI 49.1, 65.0; N= 147; Jung et al. 2020), and a single study also addressed the 6-11.9m window (17.7%; 95% CI 10.8, 25.9; N=96; Phua et al. 2005).

**Figure 2.**
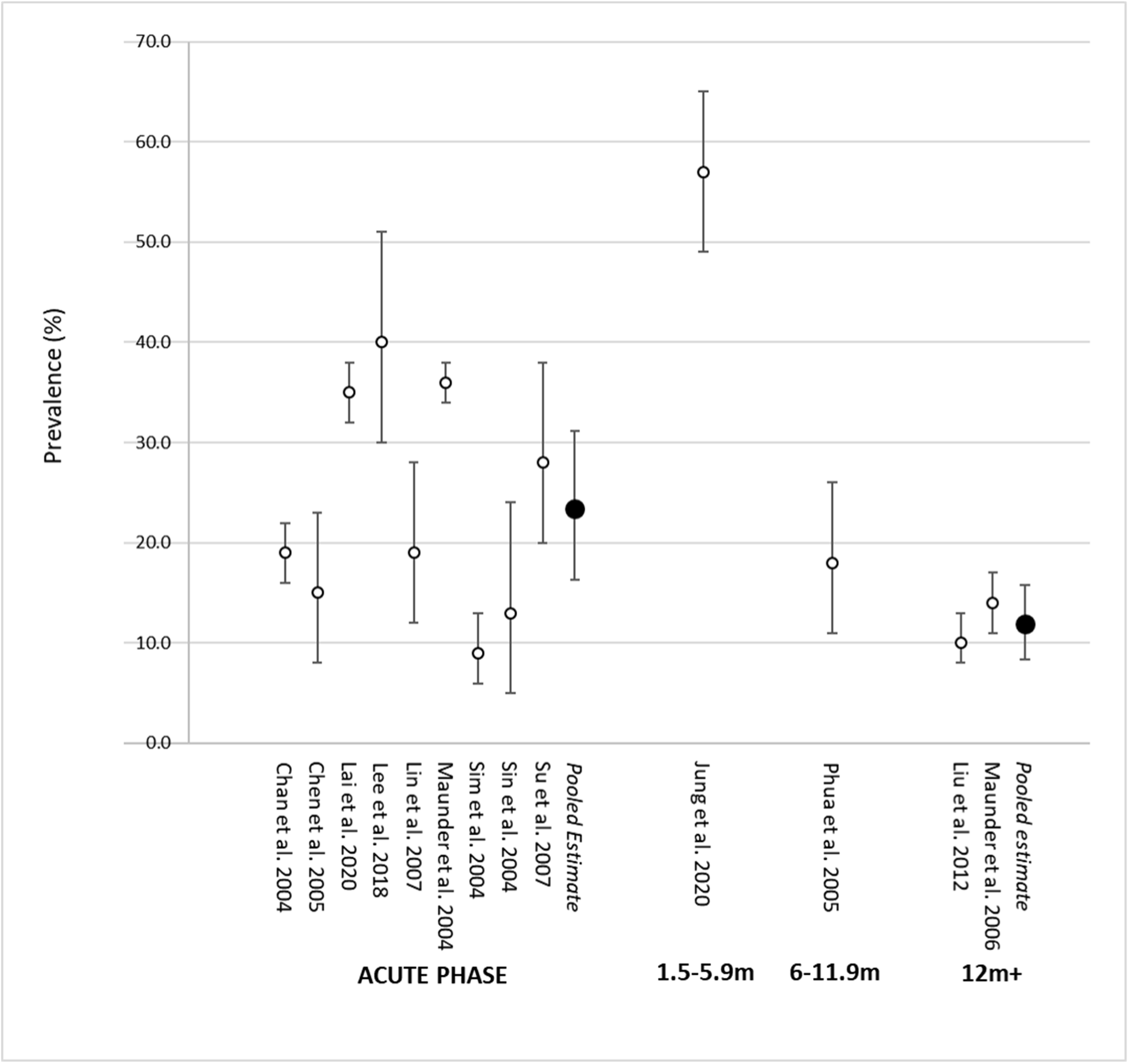
Forest plot showing prevalence of PTSS by time window.

The pooled estimate for the period 12 months onwards was 11.9% (95% CI 8.4, 15.8; k=2; N=1136). The pooled studies had a large degree of heterogeneity (Q[1] = 3.89, p<.05; I^2^=74.3%). One of these studies^14^ also conducted structured clinical interviews for PTSD with a sub-set of HCWs in Toronto (139 of 587; 24%). Two HCWs met criteria for current PTSD, with one identifying the SARS experience as the most severe traumatic event.

**Prevalence of anxiety**. One study reported the prevalence of clinically significant anxiety in the acute phase, as measured by the GAD-7^15^ (12.3%; 95% CI 10.5, 14.1; N=1257; Lai et al. 2020).

**Prevalence of depression**. Forest plots for each time window are displayed in Supplementary Material 5. The pooled estimate for depression in the acute phase was 20.2% (95% CI 9.5, 33.7; k=2; N=1359). The pooled studies had a large degree of heterogeneity (Q[1] = 9.26, p<0.003; I^2^=89.2%). A further study considered the prevalence of depression in the 12 month onwards window (8.7%; 95% CI 6.5, 11.3; N=549; Liu et al. 2012). In their study that utilised structured interviews for psychiatric disorders, Lancee and colleagues^14^ found that there was one new case with a major depressive episode, among 93 HCWs who reported no pre-SARS mental health disorders.

**Prevalence of general psychiatric screening cases**. Forest plots for each time window are displayed in Figure 3. The pooled estimate for the prevalence of positive cases on general psychiatric screening instruments in the acute phase was 34.1% (95% CI 18.7, 51.4; k=8, N=3971). The pooled studies had a large degree of heterogeneity (Q[7] = 1199.28, p<0.0001; I^2^=99.1%).

**Figure 3.**
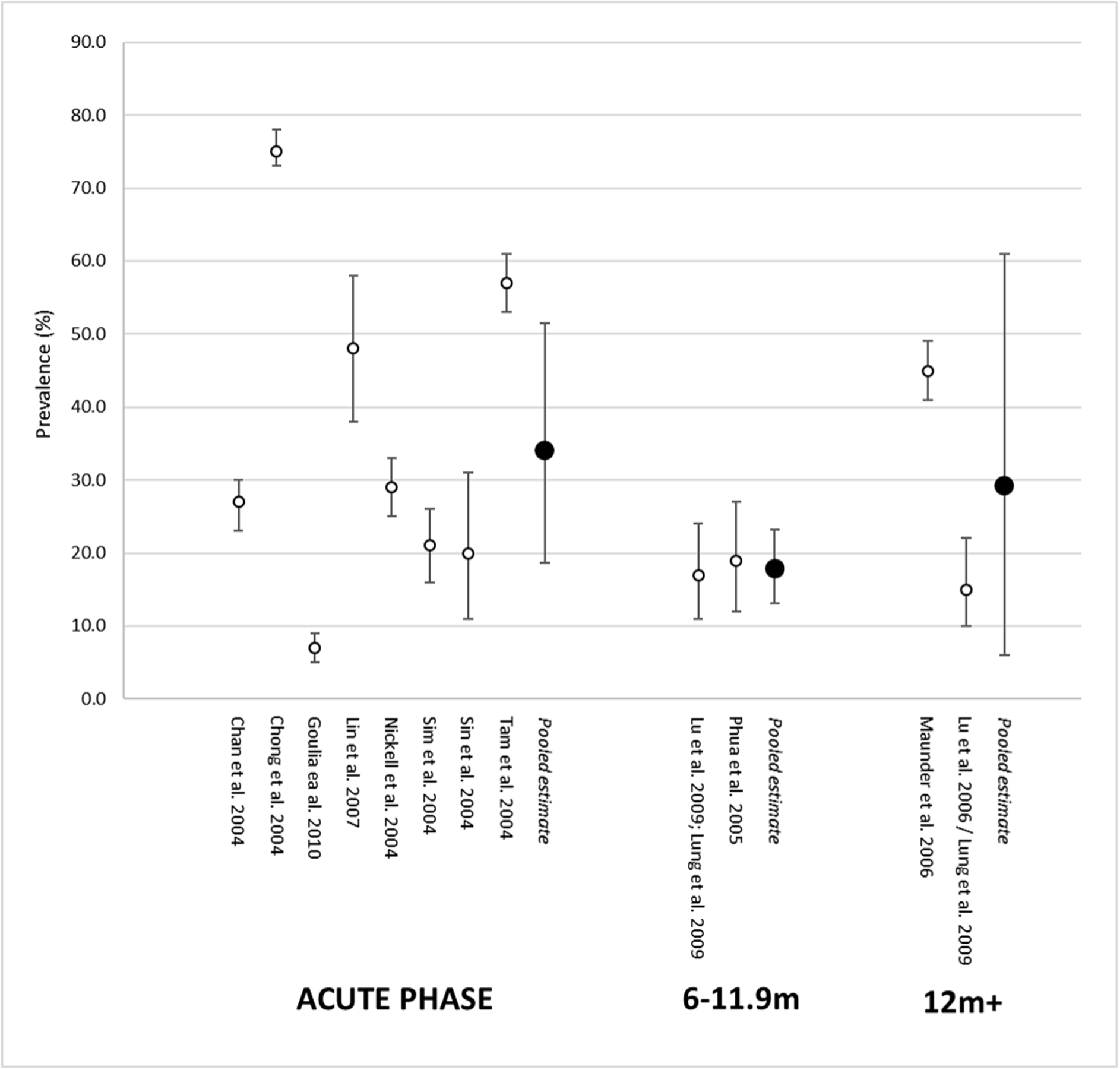
Forest plot showing prevalence of General Psychiatric Screening by time window.

For the 6 to 12 month window the pooled estimate was 17.9% (95% CI 13.1, 23.2; k=2; N=223), with no significant heterogeneity (Q[1] = 0.08, p= 0.78; I^2^=0.0%). For the 12 months onwards window, the pooled estimate was 29.3% (95% CI 6.0, 61.0; k=2; N=710). These studies were associated with a large degree of heterogeneity (Q[1]=44.60, p<.0001; I^2^=97.8%). One of these studies^13^ used a relatively low threshold for indicating caseness, possibly inflating the numbers that screened positive (44.9%); when applying the same criteria to a hospital not affected by the SARS epidemic, the authors found a large proportion (30.2%) scored above this threshold.

### Exploratory sub-group analyses in the acute phase

Given the significant heterogeneity present for most of the meta-analyses conducted, exploratory sub-group analyses were undertaken to see if more consistent findings might be discernible. Such analyses were only possible in relation to the acute phase. Moderator analyses were not undertaken given how few studies were available.

First, meta-analyses were undertaken that included only those studies which addressed the SARS pandemic (the bulk of the retrieved literature). The SARS-only prevalence meta-analysis for acute PTSS yielded a pooled prevalence estimate of 19.7% (95% CI 13.1, 27.4; k=7; N= 2813), only slightly less than the estimate for all studies. There remained a large degree of heterogeneity (Q[6] = 159.51, p<.0001; I^2^ = 93.8%). The SARS-only prevalence meta-analysis for acute general psychiatric screening cases yielded a pooled prevalence estimate of 39.1% (95% CI 23.9, 55.6; k=7; N=3502), a slightly higher than figure than that obtained for all studies. Again, this did not improve heterogeneity, which remained large (Q[6])=749.37, p.0001; I^2^=99.8%).

Second, meta-analyses that used only the same measure were undertaken. For the IES (PTSS)^16^, a point estimate of 21.0% was obtained (95% CI 11.7, 32.0; k=4; N= 2351; Q[3]=87.25, p<.0001; I^2^=95.7%). For the IES-R^17^ a point estimate of 26.6% was obtained (95% CI 9.4, 48.7; k=3; N=1611; Q[2]=97.67, p<.0001; I^2^=97.9%). For the GHQ-28^18^ a point estimate of 17.7 (95% CI 9.3, 28.1; k=4; N=1462; Q[3]=87.60, p<.0001; I^2^=94.9%). For each analysis there remained a large degree of heterogeneity.

It is noteworthy that even when restricting analyses to a single measure (e.g. IES, IES-R, GHQ-28), different cut-off scores were used to denote caseness. For example, if the point estimate GHQ-28 in the acute phase was restricted to studies that used a cut-off score of 5, a point estimate of 23.4% was obtained (95% CI 18.6, 28.5; k=3; N=993) that did not have significant heterogeneity (Q[2]=4.88, p=0.087; I^2^=58.7%).

## Discussion

The studies identified in this rapid systematic review and meta-analysis predominantly addressed clinically significant PTSS and general psychiatric caseness in HCW in the acute phase, i.e. during and immediately after a pandemic. Fewer studies addressed longer-term follow up. The majority of the studies considered the SARS pandemic. Our findings suggest that a significant minority of HCWs met threshold for clinically significant PTSS and general psychiatric caseness in the acute phase.

The limited data from follow up studies suggested that rates of PTSS declined in the months following a pandemic. Only two studies addressed PTSS rates more than 12 months post-pandemic and the results demonstrated significant heterogeneity. However, they both were large (>500 participants) and yielded a reasonably precise estimate of PTSS prevalence (95% CI 8.4-15.8%), that was markedly lower than the (albeit more imprecise) estimate for the acute phase. For general psychiatric screening the picture was not clear. While two studies suggested an improvement by the time of the 6-12 month post-pandemic window, there was considerable heterogeneity at the time on the 12 months onwards timeframe. We would highlight the potential contribution to between-study heterogeneity of using different cut-offs in screening instruments. The one study to use structured interview assessments at follow-up found very low rates of psychiatric disorder that might be directly attributable to HCW experiences during a pandemic.

We found that PTSS were elevated during the acute phase and at 12 months, similar to existing populations of at-risk health workers such as rescue workers^19^, paramedics^20^, and HCWs in general^21^ who report higher levels of PTSS than the general population. Nonetheless, whilst there were limited data pertaining the course of clinically-significant PTSS, our findings are broadly consistent with the existing literature suggesting that natural recovery is common in trauma exposed individuals^22 23^. However, it is likely that the COVID may have a longer, ongoing acute phase than in those studies reviewed such staff may have longer exposure to stress whilst experiencing PTSS. Furthermore, it is possible that COVID-19 represents a degree of threat more serious than that from previous pandemics due to factors including lack of personal protective equipment and impaired systemic resilience factors related to social distancing. As such, our findings may provide an under-estimate. Whilst these reactions are considered normal, it is imperative to consider how best to support staff during the ongoing crisis, including how to detect persistent PTSD early.

This review highlights the need for urgent research to include more extensive follow up and look to disorders other than PTSS; depression in particular has received limited attention. While screening instruments for outcomes like PTSS and depression have obvious benefits in terms of cost-effectiveness, they may also miss key aspects of HCW experience and may fail to consider the impact of other factors, e.g. prior or non-healthcare traumatic experiences. More detailed assessment (e.g. using structured interviews), similar to that undertaken by Lancee and colleagues^14^ may be warranted. It was beyond the scope of the present rapid review to identify risk factors for mental health disorders but we would strongly advocate pursuing such research.

This study is strengthened by its inclusion of studies addressing clinically similar situations, its detailed coverage of methodological issues and its *a priori* definition of time windows. The study is limited by the limited available data, and the relative narrowness of outcomes the literature has addressed.

## Conclusion

There is evidence that HCWs working in pandemics are at increased risk of a range of adverse mental health outcomes. Immediate research is needed to understand the effects of psychological stress and trauma on HCWs during COVID and how best to support HCWs during and after the pandemic.

## Data Availability

Data reported in manuscript.

## Author contributions

SA, RB, JB, TC, SP, and GS designed the study, completed the literature search and contributed to the writing of the manuscript. MB contributed to the design of the study and the writing of the manuscript. RMS designed the study, contributed to the plan of the literature search, undertook the synthesis, produced the forest plots, and contributed to the writing of the manuscript.

